# Multi-Omics Assessment of Genetic Risk for Celiac Disease in Down Syndrome

**DOI:** 10.1101/2022.09.27.22280436

**Authors:** Marisa Gallant Stahl, Jessica R Shaw, Neetha Paul Eduthan, Angela L Rachubinski, Keith P Smith, Belinda Enriquez Estrada, Matthew D Galbraith, Ronald J Sokol, Sameer Chavan, Laura Ann Leaton, Katherine M Kichula, Paul J Norman, Jill M Norris, Edwin Liu, Joaquin M Espinosa

## Abstract

**Objectives:** Individuals with Down syndrome (DS) display high risk of celiac disease (CD), but the mechanisms underlying this increased susceptibility await elucidation. Here, we examined the prevalence of HLA genotypes associated with CD risk in the general population and tested a previously developed genetic risk score (GRS) for CD in people with DS.

**Methods:** HLA genotypes were obtained for 204 individuals with DS in the Human Trisome Project cohort study, of whom 9% had CD. We compared HLA genotype frequencies in those with and without CD against frequencies observed in the general population. CD permissive HLA haplotypes explored were DQ2.5, DQ2.2, DQ8.1, and DQ7.5. We also analyzed 38 non-HLA-DQ alleles used to generate the CD GRS.

**Results:** Frequencies of risk genotypes were different for CD in DS versus CD in the general population. For example, we observed lower frequency of DQ2.5/DQ2.5 and higher prevalence of DQ7.5/X and X/X in CD in DS. Although GRS values were significantly increased in those with CD and DS, their predictive power was decreased relative to the general population. Transcriptome analysis revealed dysregulated expression of many genes composing the GRS in DS. Proteomics analysis showed that GRS values correlate with elevation of specific immune factors in DS.

**Conclusions:** The genetic risk profile of CD in DS is different relative to the general population, which is likely due to dysregulation of immune pathways in DS. Larger studies are needed to elucidate pathogenic mechanisms and to develop a validated GRS for CD in DS.

**What is Known:**

- Celiac disease is more common in individuals with Down syndrome, but the impact of HLA risk genotypes in this population is unclear.
- A celiac disease genetic risk score incorporating HLA-DQ and non-HLA SNPs has been developed with good predictive accuracy in the general population.

**What is New:**

- Individuals with DS may still develop CD even without the traditional HLA-DQ risk factors.
- A modified CD genetic risk score may be applied to individuals with DS with good accuracy and specificity.
- The immune dysregulation characteristic of DS involves dysregulated expression of many genes involved in CD etiology.

## INTRODUCTION

Down syndrome (DS) is caused by trisomy 21 (T21), the most common chromosomal anomaly, occurring in 1 in ∼700 live births ^1^. Celiac disease (CD) is more common in DS, affecting up to 10-18% of individuals with T21, representing a ∼10-fold increase relative to the general population ^2-5^. CD may be asymptomatic in DS, and even symptomatic presentation can be difficult to identify, leading to diagnostic delays ^6,7^. Despite the observation of increased prevalence, the underlying mechanisms driving this phenomenon remain unknown.

While human leukocyte antigen (HLA) predisposition represents the largest known contribution to the genetic risk of developing CD in the general population, whether or not the same HLA predispositions drive increased CD risk in DS is controversial. Some studies suggest that individuals with DS have the same HLA predispositions for CD as the general population ^3,8-11^, while others suggest that individuals with DS and CD may have unique HLA genetic risk profiles ^12-15^. Furthermore, even in the general population, HLA typing alone has poor positive predictive value ^16-18^. Many individuals with permissive HLA genotypes will not ever develop CD. Accordingly, a genetic risk score (GRS) for CD has been generated based on genetic data including both HLA and non-HLA single nucleotide polymorphisms (SNPs) from genome-wide association studies. This GRS is more discriminative of CD risk than HLA genotyping alone ^17^.

The best screening practices for CD in individuals with DS have yet to be defined, and methods for risk discrimination have yet to be examined in individuals with DS. In order to address the current gaps in knowledge, we investigated the HLA risk profile for individuals with DS and CD and explored if the GRS developed for the general population could be applied to individuals with DS.

## METHODS

### Study Participants

All participants involved in this study (n=581) were enrolled in the Crnic Institute Human Trisome Project Biobank™ (HTP). All study activities were approved by the Colorado Multiple Institutional Review Board (COMIRB) under protocol numbers 15-2170 and 20-1977. See also clinicaltrials.gov entry NCT02864108 and www.trisome.org. Samples analyzed in this study were collected between 2016 and 2020. Written informed consent was obtained from all study participants or their legal guardians. Biospecimen collection in the HTP biobank includes a blood draw, a tongue swab, and optional urine and/or stool samples. A clinical history for each participant was curated from both medical record review and a participant/family report, with medical record taking precedence in cases of discordance between the two sources. The biological datasets described in this manuscript were generated from deidentified blood-derived biospecimens and linked demographics and clinical metadata for analysis (See **Figure - Supplemental Digital Content 1**).

### Blood sample collection and processing

Peripheral blood samples were collected into PAXgene RNA and DNA Tubes (Qiagen, PAXgene RNA tubes, Cat # 762165, PAXgene DNA tubes, Cat # 761115) and BD Vacutainer K2 EDTA tubes (BD, Cat # 366643). Samples in PAXgene tubes were processed for RNA-sequencing or GWAS genotyping as described below. For the plasma-based measurements of immune markers, EDTA tubes were processed within 2 hrs of blood draw by centrifugation at 700 x g for 15 min to separate plasma, buffy coat (white blood cells, WBCs), and red blood cells (RBCs), which in turn were aliquoted, flash-frozen, and stored at -80°C. Subsequent processing was carried out as described below, with aliquots selected to minimize freeze/thaw cycles.

### Genotyping via Illumina Multi-Ethnic Global Array (MEGA) MEGA arrays

Whole genome SNP genotyping was performed using the Infinium Expanded Multi-Ethnic Genotyping Array (MEGA-EX) according to manufacturer’s instructions (Illumina LLC, San Diego, CA) as previously described ^19^.

### HLA allele imputation

We performed HLA allele imputation using the HIBAG algorithm ^20^. SNPs specific to the pre-built MEGA imputation model of HIBAG (InfiniumMEGA-Broad-HLA4-hg19) were first extracted from the whole-genome SNP data using PLINK ^21^, and duplicate SNPs removed. Imputed HLA-DQ alleles were used to infer the presence or absence of each CD-associated DQ heterodimer (DQ2.5, DQ8, DQ2.2, DQ7.5) in each participant. “X” was used to denote any heterodimer that was not one of the four listed. From the combination of heterodimers carried, we then derived the HLA-DQ genotype carried by each participant. Imputed HLA-DQ genotypes for the 19 CD cases were subsequently validated independently. *HLA* genes were targeted for DNA sequencing using a biotinylated DNA probe-based capture method ^22^, with modifications as described ^23^. Paired ends of 250 bp each were sequenced using a NovaSeq™ 6000 instrument and SP Reagent Kit (Illumina Inc, San Diego, CA). The alleles were determined from the sequence data using the consensus from three algorithms: NGSengine® 2.10.0 (GenDX, Utrecht, the Netherlands), HLA Twin™ (Omixon Biocomputing Ltd. Budapest, Hungary) and HLA*LA ^24^. There was 100% concordance between the imputed and observed genotypes.

### Genetic Risk Score (GRS)

Of 38 non-HLA-DQ SNPs in the GRS, 34 were directly genotyped on the MEGA chip. Genotypes for the remaining four SNPs were obtained through whole genome imputation via the Michigan imputation server (https://imputationserver.readthedocs.io/en/latest/prepare-your-data/). MEGA genotype data was prepared for whole genome imputation according to best practices, as recommended in documentation provided by the Michigan Imputation Server and using resources curated by Will Rayner at the Wellcome Trust Institute. Whole genome imputation was performed by the Michigan Imputation Server using the 1000 Genome Reference Panel (Phase 3, version 5) with reference population “Other/Mixed”.

All non-HLA-DQ SNPs in the genetic risk score were imputed with R^2^>0.5 and were therefore considered eligible for downstream analyses. While the published GRS used haplotype tag SNPs to infer HLA-DQ genotypes, we used the imputed HLA-DQ genotypes as described above. For one SNP located on chromosome 21 (UBASH3A rs1893592), we performed manual genotyping from raw intensity data from the MEGA chip. The Illuminaio R package was used to read in raw intensity data for this SNP. From a biplot of the allelic intensity ratio vs. the combined SNP intensity, we inferred four genotype clusters corresponding to 0, 1, 2, or 3 copies of the measured allele. We then inferred the dosage of the GRS allele, allowing up to three copies of the allele to be included in the GRS calculation. Following genomic imputation, SNPs required for the GRS calculation were extracted from imputed VCF files. Dosage data was then cleaned in R to reflect the dosage of GRS allele for each SNP. The GRS for each participant was then calculated using a log-additive model in which dosage of 15 HLA-DQ haplotype (0 or 1) and the dosages of 38 non-HLA-DQ SNPs (0, 1, 2 or 3) are multiplied by the corresponding log odds ratios as published previously ^17^ using the following formula:

GRS score = logOR_HLADQ2.5/2.5_ * dosage_HLADQ2.5/2.5_ + … logOR_HLADQn_ * dosage_HLADQn_ + logOR_SNP1_ * dosage_SNP1_ + … logOR_SNPn_ * dosage_SNPn_.

See **Table - Supplemental Digital Content 2** for HLA genotypes and **Table - Supplemental Digital Content 3** for non-HLA SNP data.

### Whole blood transcriptome analysis

Whole-blood RNA was extracted from PAXgene RNA Tubes and purified using a PAXgene Blood RNA Kit (Qiagen, Cat # 762164). RNA quality was assessed using a 2200 TapeStation system (Agilent) and quantified on a Qubit fluorometer (ThermoFisher Scientific). Globin RNA depletion, poly-A(+) RNA enrichment, and strand-specific library preparation were carried out using a GlobinClear kit (ThermoFisher Scientific, Cat # AM1980), NEBNext Poly(A) mRNA Magnetic Isolation Module, and NEBNext Ultra II Directional RNA Library Prep Kit for Illumina (New England Biolabs, Cat #s E7490 and E7760). Paired-end 150 bp sequencing was carried out on an Illumina NovaSeq 6000 instrument by Novogene Co., Ltd and delivered in FASTQ format.

### Profiling of plasma inflammatory markers using Meso Scale Discovery (MSD) assays

For each EDTA plasma sample, two technical replicates (12-25 μL depending on required dilution) were measured using the MSD multiplex immunoassay platform V-PLEX Human Biomarker 54-Plex Kit (Cat # K15248D) on a MESO QuickPlex SQ 120 instrument. Assays were performed according to manufacturer instructions and concentration values were calculated against a standard curve using provided calibrators. MSD data are reported as concentration values in picograms per milliliter of plasma and log transformed for improved visualization.

### Statistical analysis

Unless stated otherwise, all statistical analyses were performed in R (version 4.0.4).

The frequency of each HLA-DQ genotype was calculated among individuals with DS with and without CD. We qualitatively compared HLA genotype frequencies among CD cases and controls with DS to published frequencies observed in the general population ^17^.

Principal components for genetic ancestry were estimated from directly observed genotypes using PLINK 1.9 ^21^. Input to the principal components analysis calculation included only the samples eligible for inclusion in the GRS analysis, namely unrelated individuals with DS, known CD status, and complete genotypes for all genetic predictors in the GRS. The detailed methods as well as PCA plots colored by self-reported race and ethnicity are shown in **Supplemental Methods - Supplemental Digital Content 4**. The first five principal components were used as covariates in logistic regression modeling of CD cases versus controls in DS to calculate odds ratios for each SNP used in the GRS. Having derived the GRS summary metric, we evaluated its association with CD in DS and quantified its ability to predict CD in these individuals. To evaluate whether the GRS was significantly associated with CD in DS, we utilized a logistic regression model for which the outcome of CD was predicted by the GRS. From the fitted logistic regression model, we estimated a receiver operating characteristic (ROC) curve using the R package pROC (version 1.17.0.1). The Area Under the ROC (AUC) was used to quantify the predictive capacity of the GRS in individuals with DS. This was compared to the published predictive capacity of the CD GRS in the general population ^17^.

To generate the DS-GRS, the observed odds ratios in DS were used in place of the published weights in the formula described above. See **Supplementary Digital Content 2-3** for the odds ratios in DS. Differential gene expression of the included non-HLA-DQ SNPs among those in the HTP with and without DS and those with DS with and without CD was performed using R package DESeq2 (version 1.28.1) ^25^ with age, sex and sample source as covariates.

Spearman correlation analysis was used to define associations between plasma immune factors and the calculated DS-GRS using the *rcorr* function from the Hmisc package (v 4.5-0). Multiple comparisons were accounted for using the Benjamini-Hochberg (false discovery rate) method.

## RESULTS

### Individuals with Down syndrome and celiac disease display a distinct genetic risk profile

To investigate the genetic risk profile of CD in DS, we analyzed multi-omics datasets generated by the HTP. At the time of this analysis, within the larger HTP cohort, 204 participants had genomics data available generated by Illumina MEGA arrays (see **Figure - Supplemental Digital Content 1**). Nineteen of these participants had a self-reported and/or medical record diagnosis of CD (see **Table - Supplemental Digital Content 5**).

After deriving HLA haplotypes for all participants with DS, we performed a comparison of HLA frequencies in those with and without CD relative to the frequencies observed in the general population ^17,26^. For those with DS without CD, the distribution of HLA-DQ frequencies did not differ significantly with respect to the unaffected general population (**Figure 1A-B**). However, among those with CD and DS, we observed a higher frequency of traditionally low risk HLA-DQ genotypes relative to those in general population affected by CD. In fact, 11% of individuals with DS harbor traditionally low risk genotypes (DQ-7.5/7.5, DQ-7.5/X, and DQ X/X) compared to 3% of individuals in the general population (see **Table - Supplemental Digital Content 2**). Nevertheless, most (17/19) individuals with CD and DS still present with traditionally permissive genotypes. Interestingly, none of the participants with CD and DS presented DQ-2.5 homozygosity, which has been observed to be the highest risk genotype in the general population ^26,27^. It should be noted that the study may have been underpowered to detect DQ-2.5 homozygosity. Participants with CD and DS were most likely to have the DQ-2.2/X and DQ-2.5/X genotypes (21% and 26%, respectively). Participants with CD and DS also did not have the DQ-8/X genotype, which is why an odds ratio for this genotype is not included in **Figure 1B**. Next, we examined the odds of CD among the different non-HLA risk alleles included in the GRS developed with data from the general population (see **Table - Supplemental Digital Content 3**). Sixteen of the included risk alleles were associated with increased odds of CD in DS, while twenty-two were associated with decreased odds of CD in DS (**Figure 1C**). After accounting for multiple comparisons, no individual risk variant was significantly associated with CD risk in DS. On average, individuals with CD and DS display higher GRS values relative to those with DS and no CD, albeit with strong inter-individual variation (**Figure 1D**). In fact, one of the participants with CD and DS presented the lowest GRS value among the entire cohort, thus revealing the limitations of the GRS in calling CD in DS (**Figure 1D**). Overall, the published GRS yielded an AUC of 0.735 in DS (**Figure 1E**), a lesser performance relative to that observed in the general population, where it provided an AUC of 0.835 as reported by Sharp et al ^17^. With this in mind, we incorporated the observed odds ratios for each of these SNPs in DS and reweighted the GRS creating a DS-specific CD GRS (**Figure 1D**). Expectedly, the DS-GRS performed better than the published GRS ^17^ with an AUC of 0.888 (**Figure 1E**, see **Figure - Supplemental Digital Content 6**).

**Figure 1.**
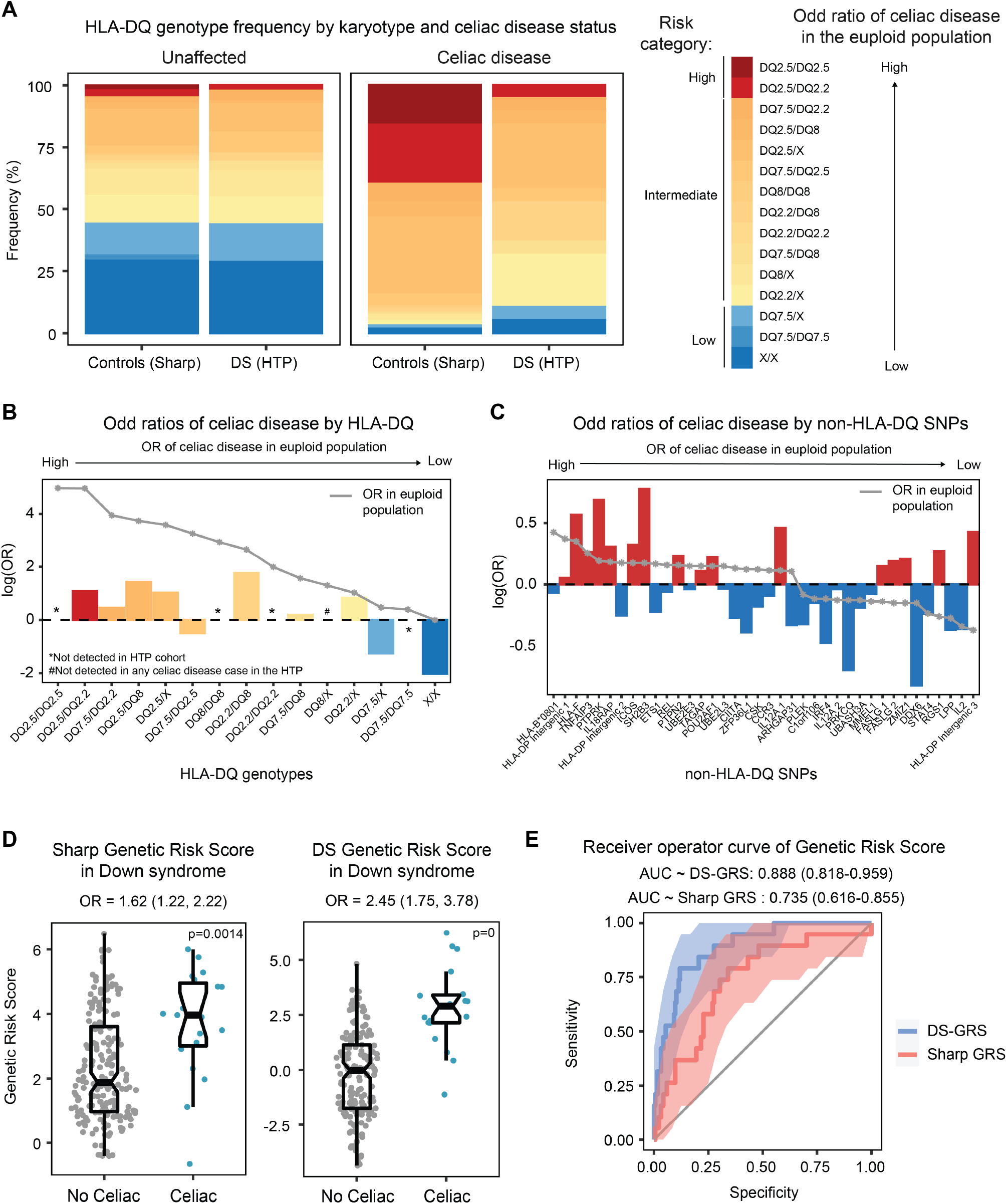
**(A)** Stacked bar plot showing the HLA-DQ frequency distribution in those with and without celiac disease and with and without Down syndrome (DS). The high-risk genotypes are highlighted in shades of red and the traditionally low-risk genotypes are highlighted in shades of blue. **(B)** Bar graphs displaying the odds of celiac disease in individuals with Down syndrome enrolled in the Human Trisome Project based on the presence of specific HLA-DQ genotypes. The gray line represents the odds ratio published in Sharp, et al. for the general population. **(C)** Bar graphs displaying the odds of celiac disease in individuals with Down syndrome enrolled in the Human Trisome Project based on the presence of specific non-HLA-DQ SNPs. The gray line represents the odds ratio published in Sharp, et al. for individuals without Down syndrome. **(D)** Sina plots displaying genetic risk score (GRS) values based on celiac disease status. The plot on the left uses the published GRS developed by Sharp et al. and the plot on the right uses the GRS reweighted with the observed odd ratios in the Human Trisome Project (DS-GRS). **(E**) Receiver operator curves of the published celiac disease GRS (red) and the DS-GRS (blue) for individuals with Down syndrome enrolled in the Human Trisome Project.

Altogether, these results indicated that increased risk of CD in DS is associated with a differential genetic risk profile, whereby risk alleles identified in the general population may have either lower or higher predictive value in DS.

### Trisomy 21 dysregulates expression of genes involved in the celiac disease risk profile

Next, we hypothesized that the differential genetic risk profile of CD in DS could be explained, in part, by effects of T21 on the expression of genes composing the GRS. We reasoned that if T21 either downregulates or upregulates mRNA expression for one or more of these genes, especially non-HLA risk genes, this would impact the behavior of the GRS in terms of predicting CD in DS. To test this, we analyzed whole blood transcriptome data generated through the HTP for individuals with and without DS (T21, n=304 versus D21, n=96), which enabled us to define the differential impacts of karyotype and CD diagnosis. Indeed, we observed that twenty of the non-HLA genes in the GRS were differentially expressed in individuals with DS, with 8 being significantly downregulated and 12 significantly upregulated (**Figure 2A-C**, see **Figure - Supplemental Digital Content 7**). Additionally, three of these non-HLA risk genes were also differentially expressed based on CD diagnosis in DS (i.e., *PTPRK, ICOS*, and *ARHGAP31*) (**Figure 2D**).

**Figure 2.**
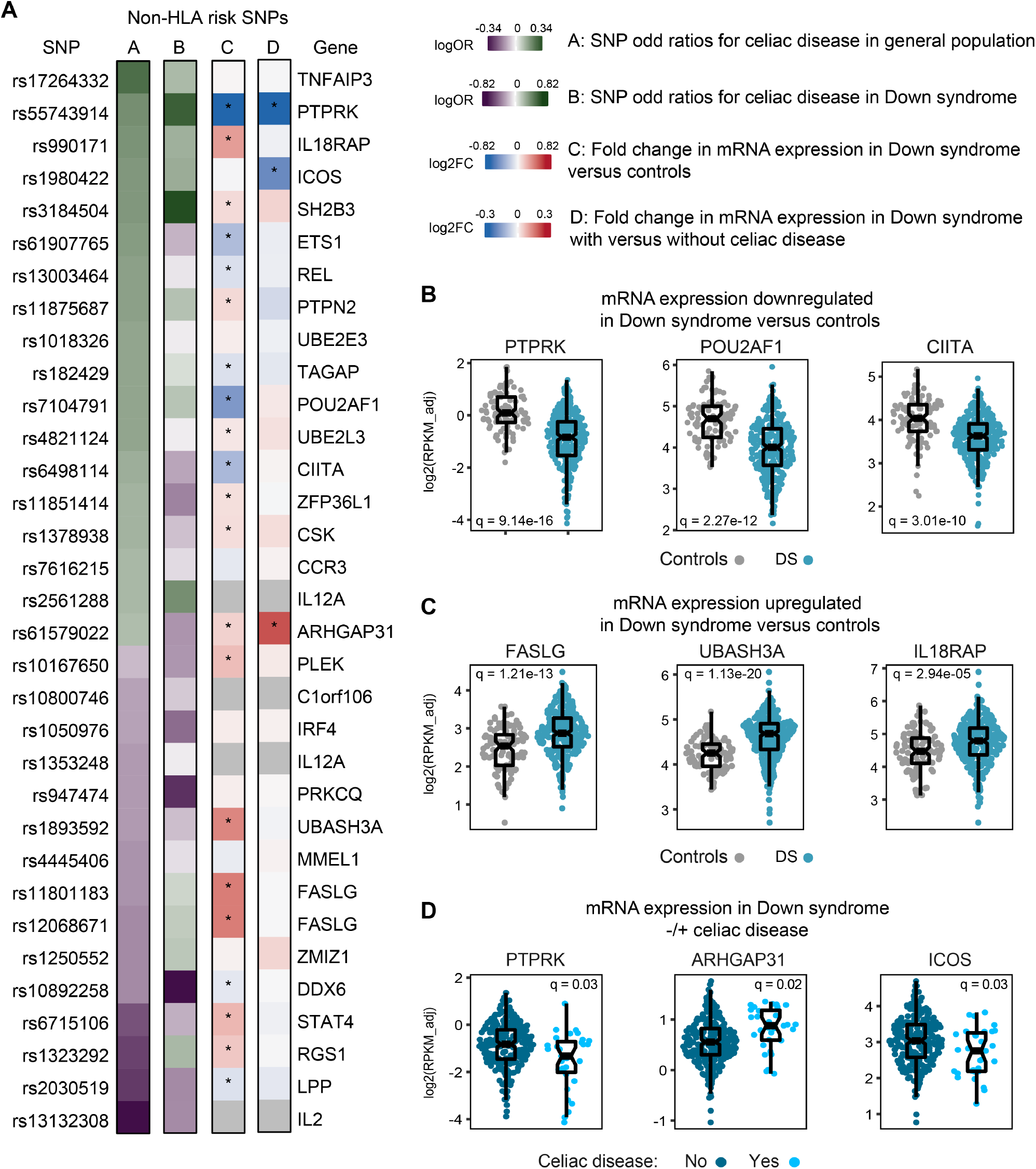
**(A)** Heat maps showing odd ratios (OR) of celiac disease (heatmaps A and B) and fold change in gene expression (heatmaps C and D). Heat map A shows ORs (log scale) of celiac disease for indicated SNPs from published data in Sharp et al for the general euploid population (controls). Heat map B shows OR of celiac disease for the same SNPs in individuals with Down syndrome in the Human Trisome Project. Heat map C shows fold change (log2 scale) in mRNA gene expression in whole blood transcriptome of individuals with T21 versus euploid controls in the Human Trisome Project. Heat map D shows fold change in gene expression in individuals with Down syndrome with versus without celiac disease in the Human Trisome Project. *Denotes a q-value <0.1 (false discovery rate 10%) Gray boxes represent SNPs for which expression data was not available. **(B-C)** Sina plots showing differences in gene expression in the whole blood transcriptome of individuals with Down syndrome versus euploid controls. **(D)** Sina plots showing differences in gene expression in the whole blood transcriptome of individuals with Down syndrome with versus without celiac disease.

Prominent examples of CD risk genes downregulated in DS include *PTPRK*, which encodes a protein tyrosine phosphatase previously associated with CD risk in the general population, and which was found to be downregulated in the mucosa of active CD compared to treated patients and unaffected controls ^28^; *POU2AF1*, a transcriptional cofactor expressed primarily in B cells ^29,30^; and *CIITA*, a transcriptional cofactor essential for major histocompatibility complex class II gene transcription ^31,32^. Other examples of downregulated genes in DS are *ETS1, REL, TAGAP, LPP* and *DDX6*. Among CD risk genes upregulated in DS, salient examples include the FAS ligand (*FASLG*), a key inducer of apoptosis ^33^; *UBASH3A*, a gene encoded on chromosome 21 involved in regulation of T cell receptor activity ^34^; and *IL18RAP*, which encodes an accessory subunit of the IL18 receptor, with mutations being associated with inflammatory bowel disease ^35,36^. Other upregulated genes in DS include *STAT4, PLEK, RGS1, ARHGAP31, PTPN2, SH2B3, CSK, ZFP36L1*, and *UBE2L3*.

Altogether, these results demonstrate that T21 affects the expression of CD risk genes, which in turn could lead to differential contributions of these genes, as well as HLA-DQ genotypes, to the genetic risk profile in DS.

### Genetic risk score associates with specific immune markers in Down syndrome

Next, we hypothesized that the genetic risk for CD in DS would correlate with differences in the inflammatory profile of individuals with DS. To test this, we defined Spearman correlations between the re-weighted DS-GRS and 54 inflammatory markers measured in matching plasma samples by MSD assays, using T21 samples only. This revealed that four immune markers were significantly positively correlated with the DS-GRS among individuals with T21: eotaxin (CCL11), eotaxin-3 (CCL26), MCP-4 (CCL13), and IL-27 (**Figure 3A-B**). Eotaxin and eotaxin-3 are chemokines that stimulate the migration of eosinophils and their role in atopic disease has been well described, but there are no previous reports of an association with CD ^37^. MCP-4 is a chemokine that has also been implicated in atopic disease and autoimmune diseases like rheumatoid arthritis, but also has not previously been associated with CD^38,39^. IL-27 is a cytokine for which expression has previously been tied to the differentiation of gliadin-specific T cells ^40,41^. These plasma chemokine and cytokine levels were also compared for those with and without DS and those with DS with and without CD with several significant differences being observed in each of these comparisons (**Figure 3C-D**). Whereas eotaxin and MCP4 are significantly elevated in the cohort with DS versus euploid controls, eotaxin 3 and IL-27 are not (**Figure 3C**). Furthermore, eotaxin, eotaxin 3, and MCP-4 are significantly elevated in individuals with DS and CD versus individuals with DS and no CD (**Figure 3D**).

**Figure 3.**
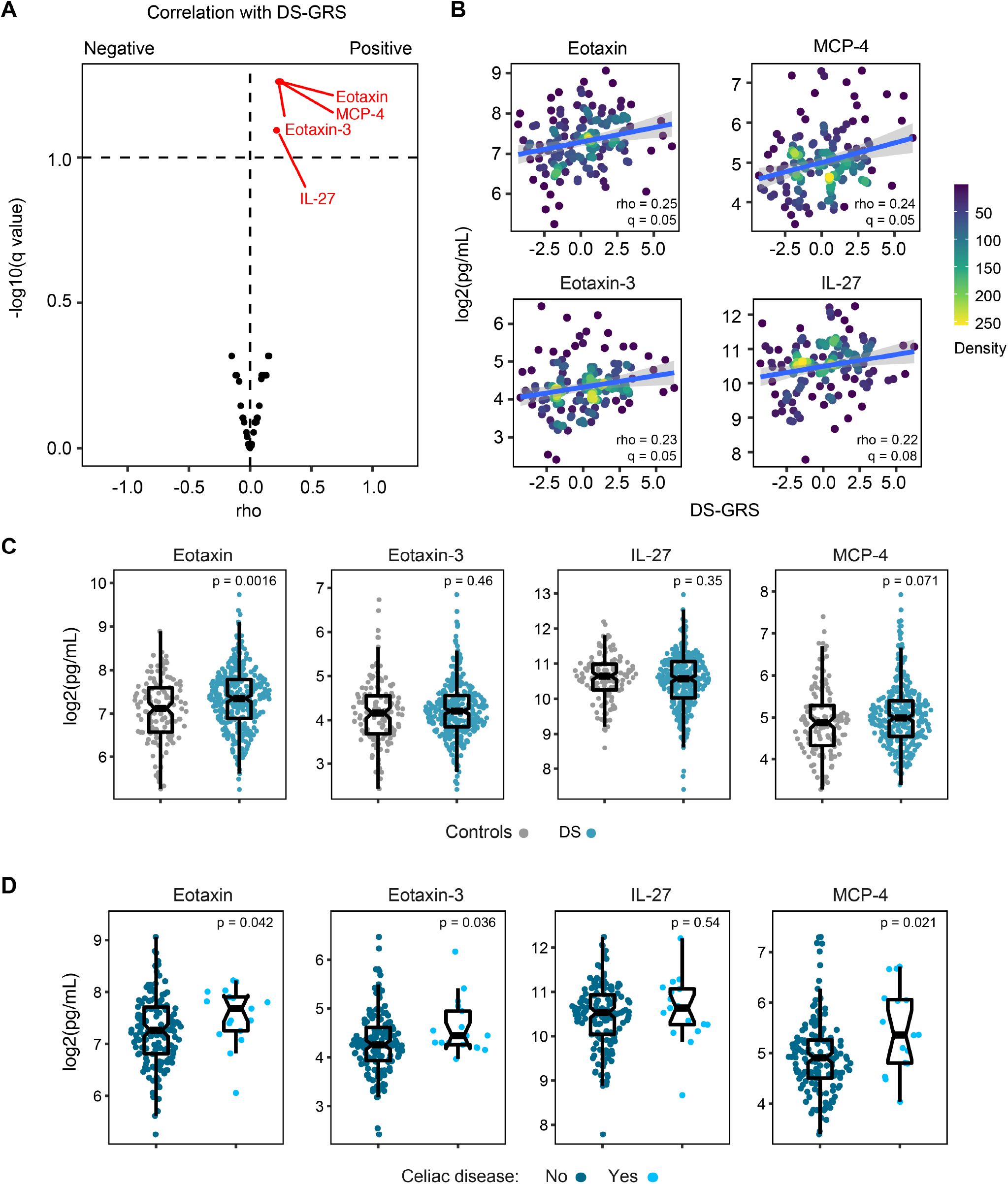
**(A)** Volcano plot displaying the correlation of inflammatory markers measured in plasma samples from individuals with Down syndrome versus their genetic risk score for celiac disease weighted based on SNP frequencies observed in Down syndrome (DS-GRS). **(B)** Scatter plots displaying correlation between plasma levels of indicated inflammatory markers and DS-GRS values in individuals with Down syndrome. Rho and q values are derived from Spearman correlation analysis. Lines indicate linear regression fit with shaded gray area displaying the 95% confidence interval. Dots are colored by density. (**C-D**) Sina plots comparing levels of indicated immune factors in individuals with Down syndrome versus euploid controls **(C)** and in individuals with Down syndrome with versus without celiac disease **(D)**. p values in Cand D calculated by Wilcoxon rank sum test.

Overall, these results suggest that the genetic risk of CD in DS is accompanied by a distinctive inflammatory milieu among those with T21.

## DISCUSSION

It is well established that individuals with DS are highly predisposed to multi-organ autoimmunity, as evidenced by high rates of autoimmune thyroid disease, autoimmune skin disorders, and CD, but the underlying mechanisms by which T21 causes these effects are not well understood. In the general population, CD risk stratification based on HLA-DQ genotype has been well characterized and is used in the clinic. Here, we demonstrate that individuals with CD and DS may not carry the traditionally high-risk HLA-DQ genotypes. Furthermore, applying a CD GRS developed for the general population that includes information on many non-HLA-DQ alleles, we found that while GRS values are indeed higher in those with CD and DS, the predictive accuracy of the GRS is decreased in DS. Clearly, a better understanding of such differential genetic risk profile for CD in DS may shed light on the increased prevalence of autoimmunity in this at-risk population more broadly, but also enhance our understanding of CD etiology and development of autoimmunity in general.

Previous studies have yielded mixed results with respect to HLA-DQ risk for CD in DS. Some studies demonstrated that individuals with DS carry the same typical HLA DQ-2.5, DQ-2.2, and DQ-8 predispositions for CD as the general population ^3,8-11^, while others demonstrated that individuals with CD and DS do not always carry these typical HLA-DQ predispositions ^12-15^. Reasons for disparate results include small sample sizes of individuals with CD and DS and different technologies for HLA-DQ typing. The largest previous study included 12 individuals with suspected CD and DS ^14^. Several studies only evaluated for DQ-2.5 and/or DQ-8 missing many of the traditionally permissive HLA-DQ genotypes such as DQ-2.2 or DQ-2.5 in the trans conformation. Our study used a well-described HLA imputation method ^20^ and confirmed these imputed HLA-DQ genotypes with Next Generation Sequencing to accurately evaluate for the presence of traditionally permissive HLA-DQ haplotypes in 19 individuals with CD and DS versus 182 controls with DS and no CD, thus becoming the largest study of CD genetic risk in DS to date. In individuals without DS, HLA-DQ genotyping is often used to rule out CD in individuals on a gluten-free diet, in individuals with inconclusive biopsy results, and in at-risk populations such as those with a family history for risk stratification and screening guidance ^42^. In fact, the negative predictive value of HLA-DQ genotyping in individuals without DS approaches 100%^43,44^. However, our results indicate that HLA-DQ genotyping may not have the same clinical application in individuals with DS and even those with traditionally low-risk HLA-DQ genotypes may still be at risk of developing CD. Therefore, these observations reveal the need for a customized approach for CD screening in DS.

To further explore the genetic profile of CD in DS, we tested a CD genetic risk score developed and validated in the general population composed of four HLA-DQ, five non-DQ HLA, and 33 non-HLA SNPs ^17^ to our DS cohort. This GRS did not perform as well in DS, which can be explained in part by the fact that the risk associated with these SNPs is different in CD in DS versus published risk for CD in the general population (**Figure 1C**). In turn, such differential risk could be linked to the fact that most of these genes were dysregulated at the mRNA level in the peripheral immune compartment of individuals with DS (**Figure 2**). Therefore, we hypothesize that the widespread dysregulation of genes involved in CD pathogenesis in DS modulates the contributions of risk SNPs identified for these genes in the general population. Given that SNPs can affect gene transcription, mRNA processing and stability, and/or protein activity, the effects of these SNPs can be attenuated or enhanced by convergent mechanisms affecting the expression and/or activities of these same genes and their products. For example, the *PTPRK* gene, located on chromosome 6, has been demonstrated to play important roles in maintenance of cell-cell junctions and has subsequently been implicated in intestinal barrier function. In the general population, individuals with active CD have been shown to have reduced expression of *PTPRK* in intestinal biopsies in line with increased intestinal barrier permeability ^28^. Our study demonstrated that *PTPRK* expression was decreased in individuals with DS compared to the general population and that expression was further reduced in individuals with DS with versus without CD, all of which may contribute to different frequencies of *PTPRK* SNPs in DS. Given that all these risk SNPs affect genes in a small number of interconnected pathways (**Figure 4**), it is difficult to predict how differences in mRNA expression may impact on SNP frequency. Furthermore, we should note that our transcriptome analysis employed whole blood RNA, which may not reflect gene expression changes in the gut epithelium or other relevant tissues.

**Figure 4.**
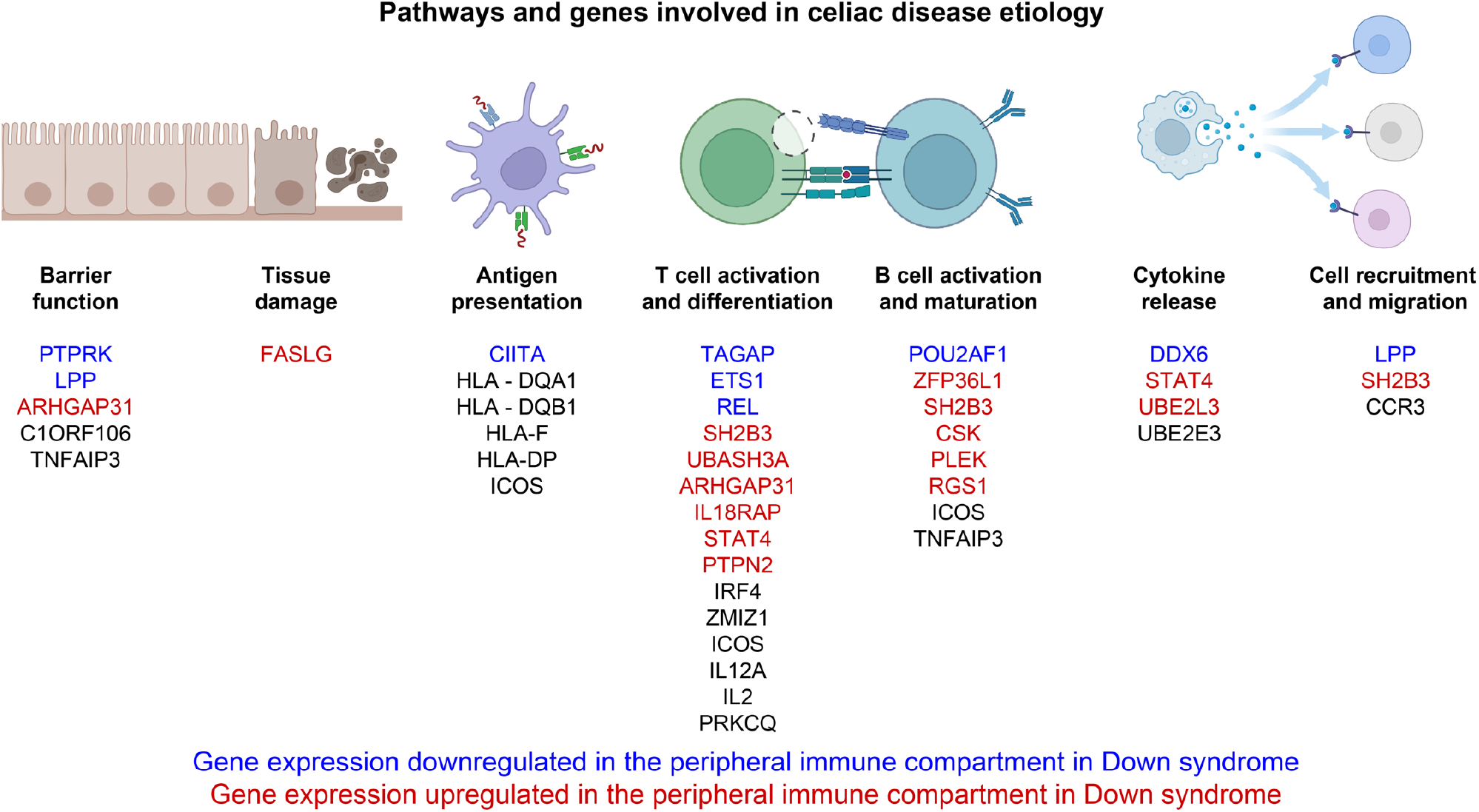
Graphical abstract displaying key pathways of celiac disease pathogenesis and the impact of trisomy 21 on the expression of genes involved in each pathway. Composed with graphic elements from BioRender.

Noteworthy, individuals with DS display chronic activation of the interferon (IFN) response, a key aspect of the innate immune system, which could contribute to their increased risk of autoimmunity, including CD. Indeed, many of the pathways involved in CD pathogenesis (**Figure 4**) are affected by IFN signaling. Such IFN hyperactivity can be explained by the fact that four of the six IFN receptors are encoded on chr21 and widely overexpressed throughout the immune cell lineages of individuals with DS ^45-48^. Given that the IFN response employs JAK1-2 kinases for signal transduction, these findings justify the ongoing testing of JAK inhibition to decrease the burden of autoimmunity in this population (NCT04246372) ^49-51^.

While these findings are interesting, several limitations must be accounted for when interpreting these results. Our study included 19 individuals with CD and DS, which is larger than previous studies, but an even larger sample size would be needed to define statistical significance for many of our observations. Another limitation is the broad clinical inclusion criteria for CD diagnosis. Four individuals only had self-reported diagnoses of CD that were unable to be confirmed in the medical record. Of the ones that were confirmed in the medical record, several individuals were diagnosed based on positive antibody results and did not receive a confirmatory intestinal biopsy. This could lead to misclassification error and future studies should include individuals with deeply annotated clinical data as well as multi-omics data to improve the generalizability of the results. Similarly, gluten-free diet status was not available at the time of the blood draw for most participants. It is known that gene expression can vary based on whether individuals are on a gluten-free diet and whether they have active CD ^28^.

Despite these limitations, our study has the largest sample size to date of individuals with CD and DS and is the first to have extensive multi-omics data available for this population. In conclusion, we have demonstrated that individuals with CD and DS have a differential genetic risk profile relative to CD in the general population that also corresponds with differences in gene expression.

## Supporting information

Digital Supplemental Content 5 - Cohorts characteristics

## Data Availability

All data produced in the present study are available upon reasonable request to the authors

## Abbreviations

(CD): Celiac Disease
(D21): Disomy 21
(DS): Down Syndrome
(GRS): Genetic Risk Score
(HLA): Human Leukocyte Antigen
(IFN): Interferon
(T21): Trisomy 21

## Author contributions

**Marisa Gallant Stahl**: conception of the work, data interpretation, funding acquisition, drafting the manuscript, final approval of the version to be published.

**Jessica R. Shaw**: data analysis and interpretation, critical revision of the manuscript, final approval of the version to be published.

**Neetha Paul Eduthan**: data analysis and interpretation, critical revision of the manuscript, final approval of the version to be published.

**Angela L. Rachubinski**: IRB protocol writing and oversight, participant recruitment and consent, biospecimen acquisition, data acquisition, critical revision of the manuscript, final approval of the version to be published.

**Keith P. Smith**: biospecimen acquisition and processing, critical revision of the manuscript, final approval of the version to be published.

**Belinda Enriquez Estrada**: IRB protocol oversight, participant recruitment and consent, biospecimen acquisition, data acquisition, critical revision of the manuscript, final approval of the version to be published.

**Matthew D. Galbraith**: data analysis and interpretation, critical revision of the manuscript, final approval of the version to be published.

**Ronald J. Sokol**: conception of the work, data interpretation, funding acquisition, critical revision of the manuscript, final approval of the version to be published.

**Sameer Chavan**: data acquisition and analysis, critical revision of the manuscript, final approval of the version to be published.

**Laura A. Leaton**: Data acquisition and analysis, critical revision of the manuscript, final approval of the version to be published.

**Katherine M. Kichula:** Data acquisition and analysis, critical revision of the manuscript, final approval of the version to be published.

**Paul J. Norman**: data acquisition and analysis, critical revision of the manuscript, final approval of the version to be published.

**Jill M. Norris**: Conception of the work, data interpretation, critical revision of the manuscript, final approval of the version to be published.

**Edwin Liu**: Conception of the work, data interpretation, critical revision of the manuscript, final approval of the version to be published

**Joaquin M. Espinosa**: Conception of the work, IRB protocol writing and oversight, participant recruitment and consent, data generation, analysis and interpretation, critical revision of the manuscript, final approval of the version to be published.

All authors agree to be accountable for all aspects of the work in ensuring that questions related to the accuracy or integrity of any part of the work are appropriately investigated and resolved.

## Acknowledgements

We would like to thank Monica Campbell, Tonya Brunetti, and the Colorado Anschutz Research Genetics Organization (CARGO) team for their assistance with preparation and analysis of the MEGA chip samples. We thank Dr. Kim Jordan at her team at the Human Immune Monitoring Shared Resource for outstanding service in generation of the immune marker dataset. We are also grateful to the Colorado Translational and Sciences Institute and the Colorado Multiple Institutional Review Board for assistance in all clinical research projects involving the Crnic Institute. Special thanks to Michelle Sie Whitten, the team at the Global Down Syndrome Foundation and Dr. John Reilly for logistical support at multiple stages of the project. We are grateful to all participants in the Human Trisome Project as well as many team members who assisted in participant recruitment, consent, biospecimen collection and processing, as well as clinical data annotation.

## Supplemental Digital Content 1 – Figure

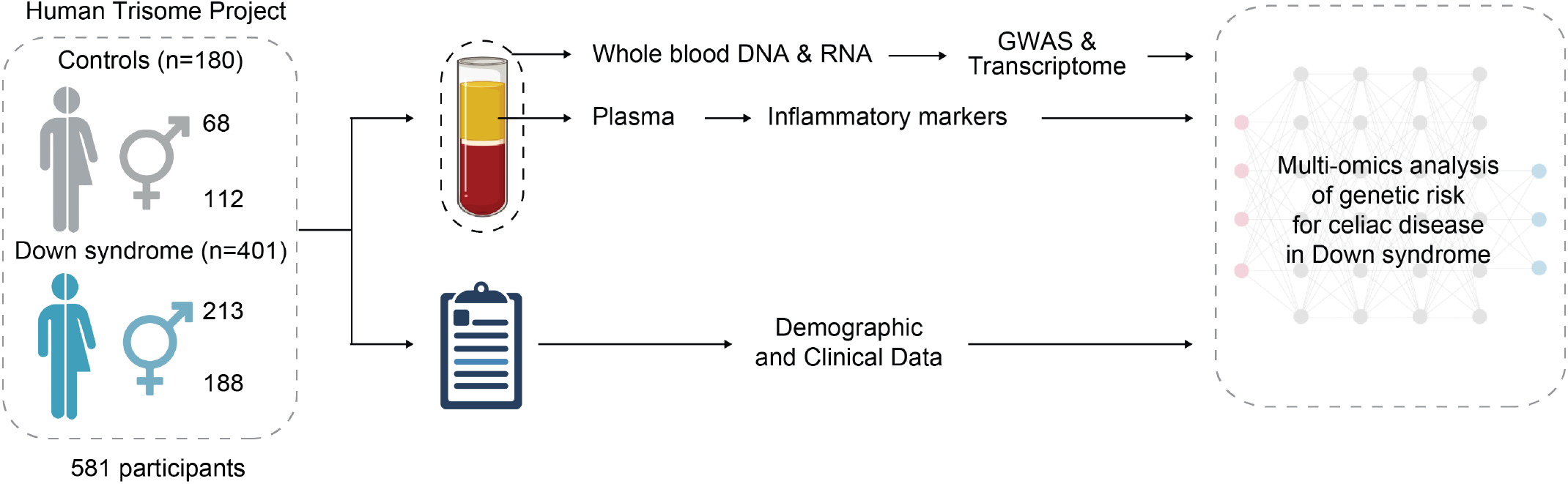

### Diagram of experimental design

Blood samples obtained from research participants in the Human Trisome Project were analyzed through a multi-omics pipeline including GWAS analysis with the Illumina MEGA array, whole blood transcriptome analysis via RNAseq, and targeted proteomics using Meso Scale Discovery assays to measure levels of inflammatory markers.

## Supplemental Digital Content 2 – Table: HLA genotype frequency by celiac status and karyotype

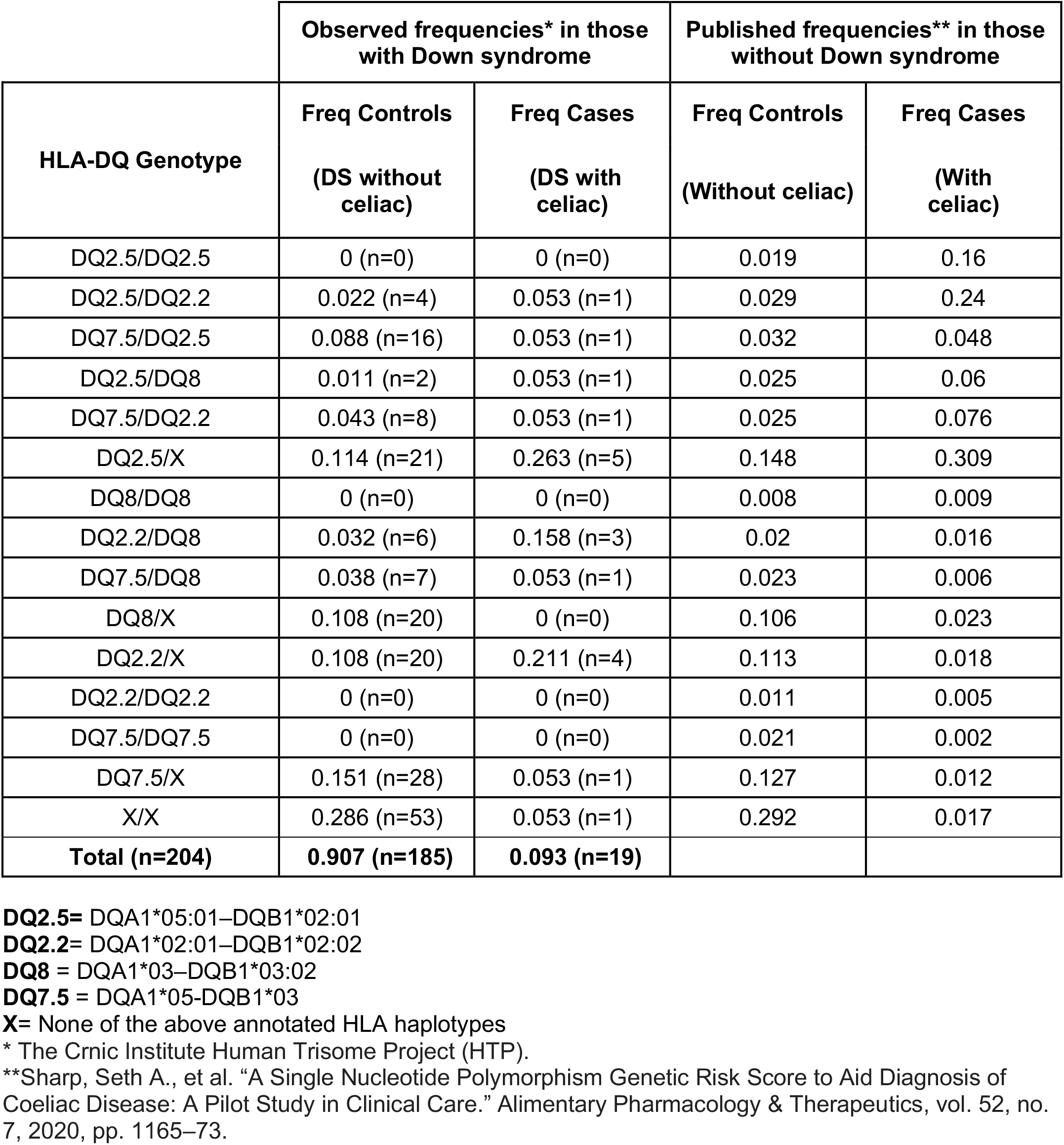

**Table:** Celiac Risk by HLA-DQ genotype.

**Table: Celiac Risk by HLA-DQ genotype**
| **HLA-DQ Genotype** | **Published OR\* in those without Down syndrome** | **Observed OR\*\*in those with Down syndrome** |
| --- | --- | --- |
| DQ2.5/DQ2.5 | 144.03 | NA |
| DQ2.5/DQ2.2 | 142.59 | 2.95 |
| DQ7.5/DQ2.2 | 51.94 | 1.58 |
| DQ2.5/DQ8 | 42.1 | 4.19 |
| DQ2.5/X | 36.23 | 2.79 |
| DQ7.5/DQ2.5 | 26.05 | 0.61 |
| DQ8/DQ8 | 18.92 | NA |
| DQ2.2/DQ8 | 14.3 | 5.84 |
| DQ2.2/DQ2.2 | 7.32 | NA |
| DQ7.5/DQ8 | 4.81 | 1.21 |
| DQ8/X | 3.67 | 0 |
| DQ2.2X | 2.77 | 2.3 |
| DQ7.5/X | 1.6 | 0.28 |
| DQ7.5/DQ7.5 | 1.48 | NA |
| X/X | 1 | 0.13 |

## Supplemental Digital Content 3 - Table: Non-HLA-DQ SNP risk of celiac disease by karyotype

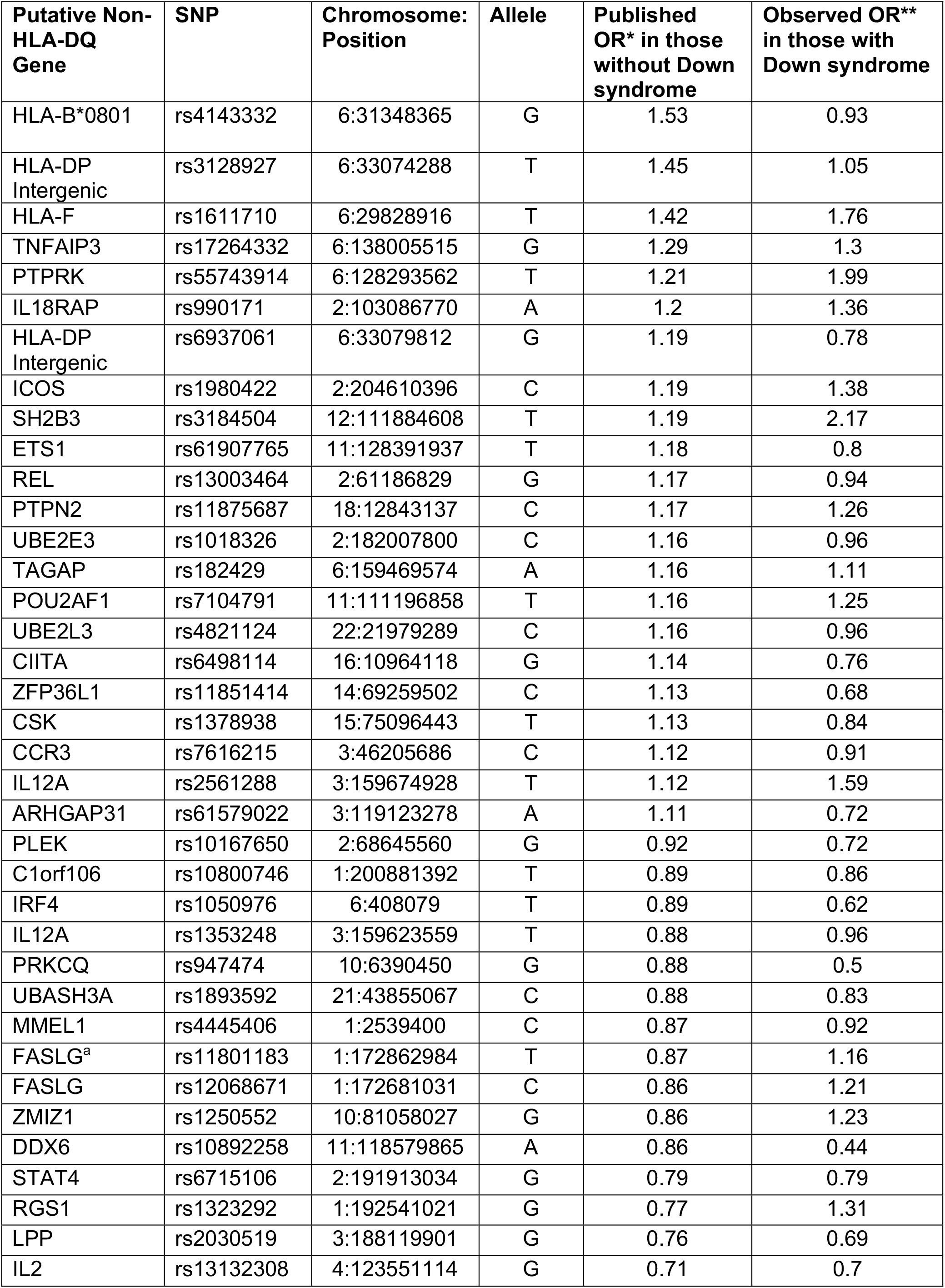
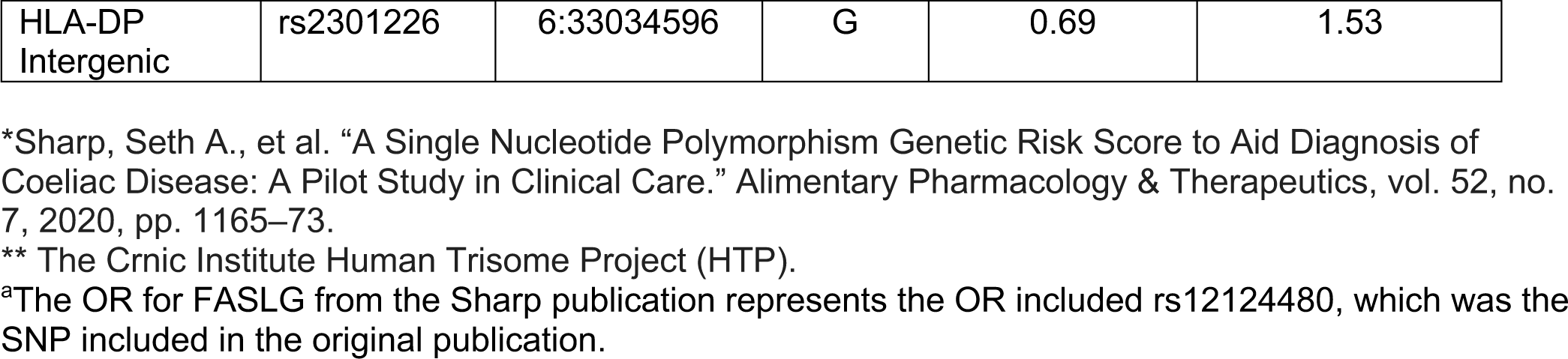

## Supplemental Digital Content 4 – Supplemental Methods

### Principal components for genetic ancestry

Excluded from input to principal components analysis calculation were chromosome 21, sex chromosomes, mitochondrial variants, variants with unknown chromosomal mapping, and the specific non-HLA-DQ SNPs used in the GRS. Also excluded from the principal components analysis calculation were ancestry-specific regions which may confound the analysis ^1,2^ including chr8:8135000-1200000, chr17: 40900000-45000000, and the HLA region plus or minus 200 kb (chr6:28277797-33648354). Because HLA alleles have been demonstrated to strongly associate with variants up to 200 kb outside of the formal boundaries of the HLA region ^3,4^, we excluded variants within the HLA region as well as variants located within 200 kb of the HLA region boundaries. We further filtered input variants to common variants with a minor allele frequency >5% (Plink --maf 0.05), sample call rate > 95% (--mind 0.05), and variant call rate > 98% (Plink --geno 0.02). Lastly, we performed SNP pruning to identify a subset of pairwise independent variants with a maximum pairwise correlation of 0.20 (Plink -- indeppairwise 50 5 0.2). The first 5 principal components (PCs) for ancestry were used as covariates in single-variant logistic regression models to account for population stratification (**Supplemental Methods Figure 1**).

**Supplemental Methods Figure 1.**
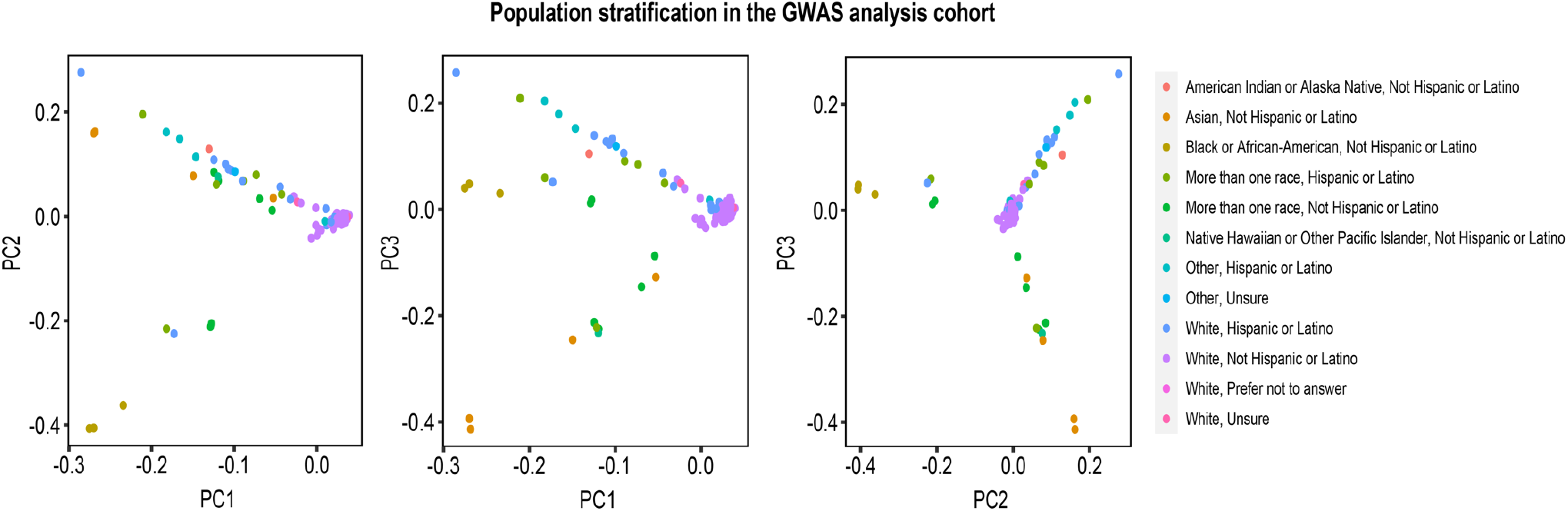
Principal component analysis plots displaying the population stratification by race and ethnicity among participants in the GWAS study of the Human Trisome Project.

### SNP calling for UBASH3A

In order to identify the probe identifier corresponding to the UBASH3A SNP of interest (rs1893592), we downloaded the MEGA product manifest from the Illumina website: https://webdata.illumina.com/downloads/productfiles/multiethnicglobal-8/v1-0/infinium-multi-ethnic-global-8-d1-csv.zip. Using the illuminaio R package, we read the raw intensity data into R and extracted a matrix of data from each idat file. We then mapped the row names of the raw intensity data to the AddressA_ID column of the Illumina product manifest to identify that AddressA_ID of ‘29800833’ corresponded to the chr21 SNP of interest, UBASH3A^rs1893592^. We identified the TOP/BOT (A/B) alleles for the probe using the strand report for the MEGA product provided on the Illumina website: https://webdata.illumina.com/downloads/productfiles/multiethnicglobal-8/v1-0/multi-ethnic-global-8-d1-strand-report.zip. Having identified the probe specific to the SNP of interest and the A and B alleles for that probe, we next derived the Combined SNP intensity (R) and the Allelic Intensity Ratio (*θ*) using the equations: **R** = Mean intensity_Red_ + Mean intensity_Grn_, and 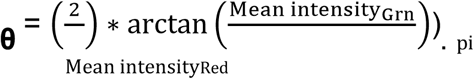

We then produced scatterplots of *θ* versus R and visually identified thresholds of *θ* that clearly separated the samples into four genotype clusters (**Supplemental Methods Figure 2**). We then used the identified theta thresholds to manually assign a genotype for each sample.

**Supplemental Methods Figure 2.**
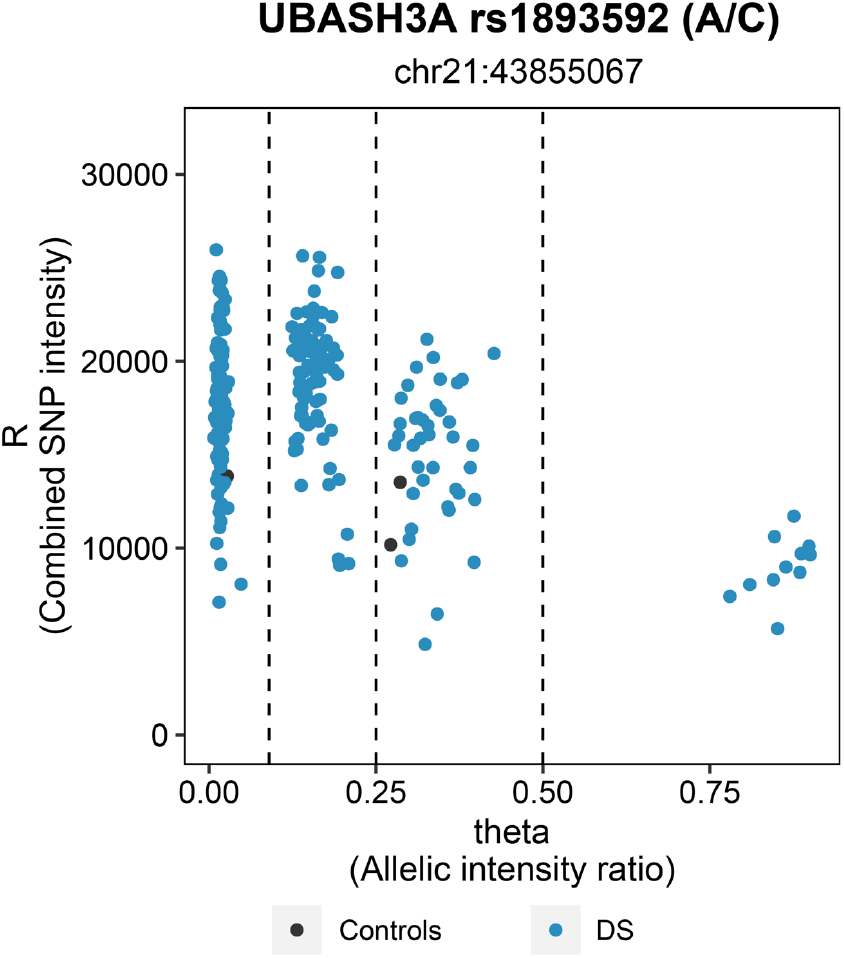
Scatter plot demonstrating the genotype assignment of the SNP UBASH3A^rs1893592^, which was the only SNP located on chromosome 21 in the genetic risk score. Given its chromosomal location, this plot demonstrates how the trisomy was accounted for in the genetic risk score.

## Supplemental Digital Content 6 – Figure

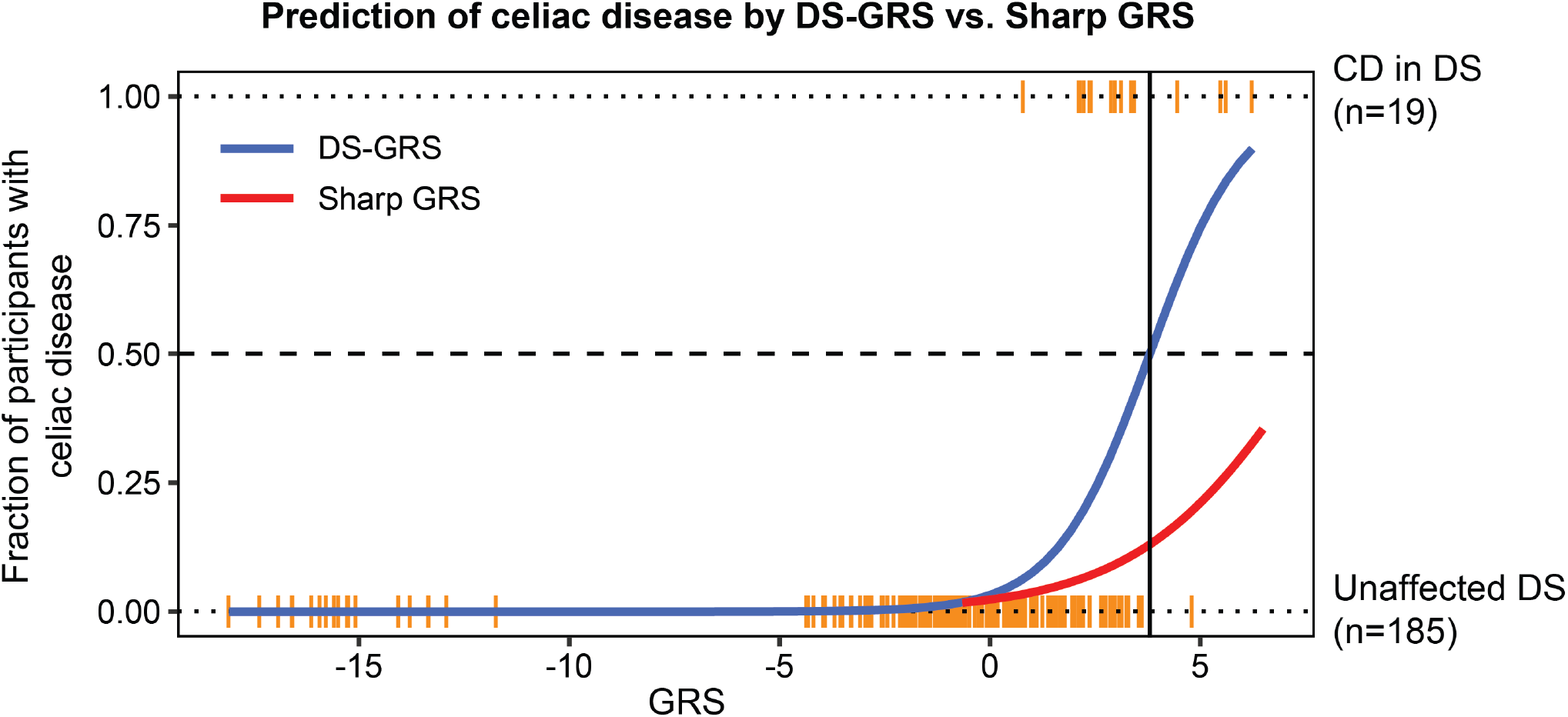

### Comparison of GRS algorithms to discriminate celiac disease in Down syndrome

GRS values were calculated as described in the Methods using a published algorithm developed with data from the general population (Sharp et al, 2020) or with a re-weighted algorithm (DS-GRS) using data from individuals with Down syndrome in the Human Trisome Project. Plot displays the comparative predictive power of the two versions of the GRS. The vertical black line indicates the value at which the DS-GRS correctly identifies 50% of celiac disease cases.

## Supplemental Digital Content 7 – Figure

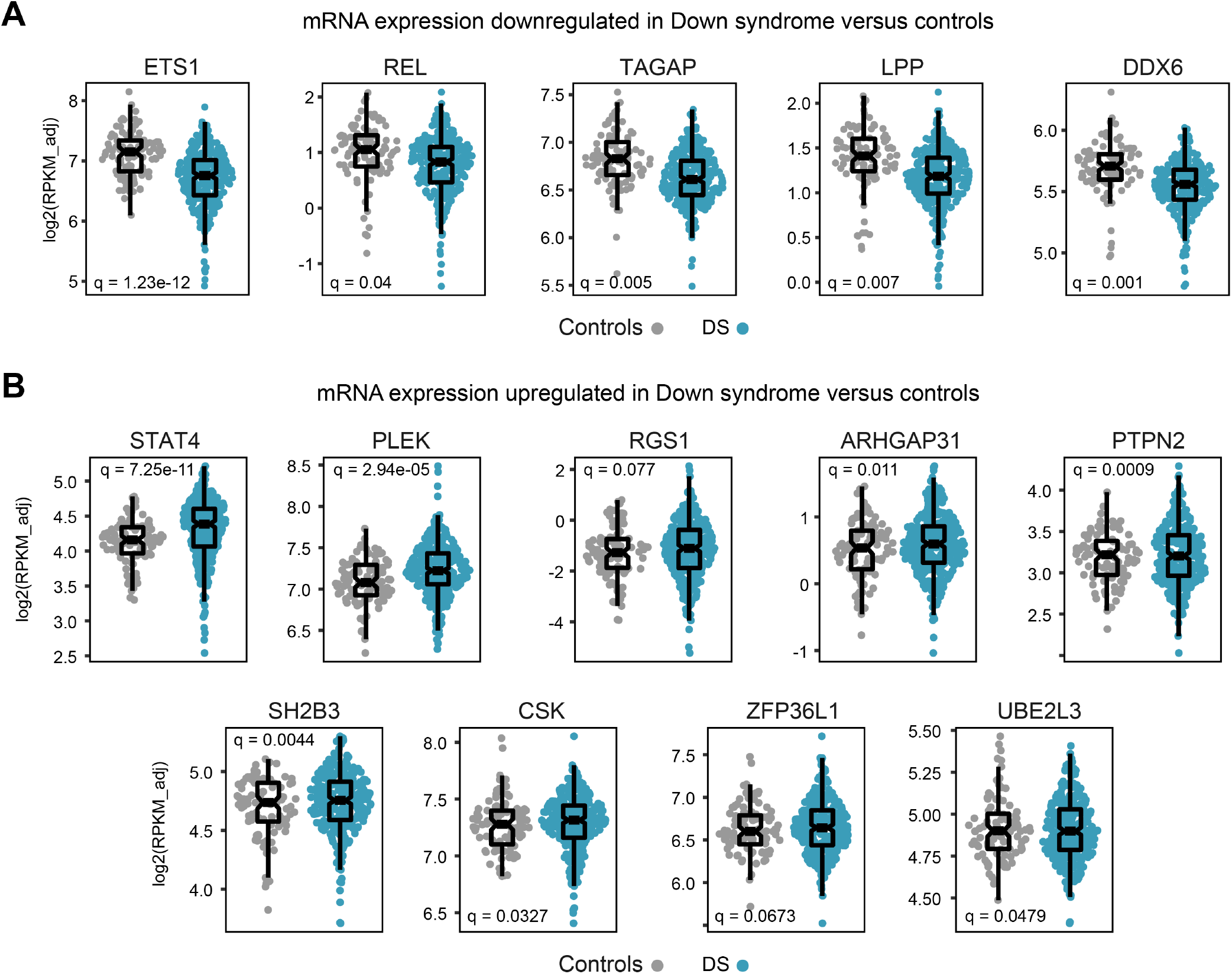

### Whole blood transcriptome analysis reveals dysregulation of multiple genes involved in the celiac disease genetic risk score

Sina plots displaying the expression of mRNAs that are either significantly downregulated (A) or upregulated (B) in individuals with Down syndrome versus euploid controls in the Human Trisome Project. Differential expression was calculated using DeSeq2 software.

